# Risk factors for acne scarring in Ecuador

**DOI:** 10.1101/2023.05.03.23289452

**Authors:** Mikaela Camacho, María Isabel Viteri, Paola Yepez, Jorge Estrella Porter, Drifa Belhadi, Caroline Barnes, Jonathan Guillemot

## Abstract

**Background:** Acne is a common disease that causes a large global disease burden. The Global Burden of Skin Disease reported that in 188 countries the burden from acne as measured by disability-adjusted life years (DALYs), years lived with disability and years of life lost is greatest in Western Europe, high-income North America and Southern Latin America. This paper aims to identify risk factors for acne scarring specific to the Ecuadorian population in order to adapt the 4-ASRAT tool accordingly.

**Methods:** This was an observational prospective study. Participants were recruited to complete a survey that was developed based on the potential risk factors for acne scarring and had facial photographs taken. To determine risk factors and their respective weighting, a logistic regression was performed.

**Results:** The study included 404 participants. Results from univariate analyses indicated that male sex (OR=2.76 95%CI [1.72; 4.43]), severe or very severe acne scarring (OR=4.28 95%CI [1.24; 14.79]), acne duration over 1 year (OR=1.71 95%CI [1.12; 2.60]), oily skin (OR=2.02 95%CI [1.27; 3.22]) and the presence of acne on the neck (OR=2.26 95%CI [1.30; 3.92]), were all significantly associated with the presence of acne scarring. Male sex (2.56 95%CI [1.58;4.17]), oily skin (1.96 95%CI [1.20;3.20]) and severe or very severe acne (3.75 95%CI [1.05;13.37]) remained significant risk factors for acne scarring in the multivariate analysis.

**Conclusion:** By identifying acne scarring risk factors and applying the tool in everyday dermatology visits, we can reduce the physical and psychological burden that acne scarring causes in the adolescent and adult population. Further research should be performed to reassess potential risk factors and complete the adaptation of the tool for the Ecuadorian population, with a larger and more representative study population.

## Introduction

Acne Vulgaris is a chronic and inflammatory skin disease that has its origin within the pilosebaceous follicles. The four main causes of this disease are sebum overproduction, abnormal shedding of follicular epithelium, follicular colonization by *Cutibacterium acnes* and inflammation^1^. Acne is a common disease that causes a large global disease burden. It was found globally that acne vulgaris is the eighth most common skin disease, with a prevalence of 9·38%. Also, depending on the country and age, 35% to 100% of adolescents have had acne at some point in their lives. In the United States, acne vulgaris is the most prevalent chronic disease affecting nearly 50 million people. These statistics alone show the gross severity of the epidemiology of acne^2^

The nature of acne, including its symptoms and sequelae, contributes to physical and psychological burden; especially in the most affected population, those aged 14-26 years old. Inflammatory acne sequelae include post-inflammatory hyperpigmentation and permanent or temporary scarring. The Global Burden of Skin Disease reported that in 188 countries the burden from acne as measured by disability-adjusted life years (DALYs), years lived with disability and years of life lost is greatest in Western Europe, high-income North America and Southern Latin America. Also, acne was ranked in the top 10 after analysing the prevalence and impact of skin diseases in 187 countries. The impact of acne on health-related quality of life may result in emotional stress, significant psychosocial burden and neuropsychiatric pathologies such as anxiety, depression and suicide^3^.

A study on acne-induced scar prevalence performed in the United States, with 1972 subjects who suffered from acne, showed that 43% of these patients suffered from acne scarring. Based on the characteristics of acne, mild and moderate acne tended to form scars in 69% of the subjects; but severe or very severe acne was most likely to develop scarring^4^. Despite the negative impacts of acne vulgaris, treatment compliance is poor because patients discontinue treatment due to early improvement, perception of worsening acne and side effects^5^. Treatments that can completely resolve acne scars are not yet available and prevention and early treatment remain the primary strategy against scars. While scars cannot be completely eliminated, recent technological advances offer treatment options for patients to improve acne scarring. Such options include microdermabrasion, dermabrasion, chemical peeling, skin needling and laser resurfacing. The issue with these technological advances is that patients face challenges concerning the cost of the treatments and the time they take. The physician has to review with the patient the modalities, the number of treatment sessions based on the severity of scarring, the side effects, the discomfort that will be experienced and the cost of such sessions, all of these are immense limitations for some patients^6^. Considering treatments are complicated, we feel it is important to develop an understanding around risk factors with the goal of eventually using this information to create strategies for preventing acne scarring in the first place.

A systematic review found seventeen risk factors associated with the development of 6acne scarring: acne inflammation, gender, ethnicity, smoking, duration of acne therapy, treatment compliance, treatment fatigue, age, patient demographics, seborrhoea, body mass index, relapse after treatment, and picking and squeezing behavior^7^. The same study developed a Four-item Acne Scar Risk Assessment Tool (4-ASRAT) to help identify individuals at risk of scarring. The four risk factors found to be significantly associated with increased risk of scarring were severity of acne, family history of scarring, squeezing and picking behaviors, and duration of acne. It is however unclear how the risk factors may vary according to different regions and populations.

To our knowledge, no previous studies have explored acne-scoring risk factors in Ecuador or Latin America. This paper aims to identify risk factors for acne scarring specific to the Ecuadorian population in order to adapt the 4-ASRAT tool accordingly.

## Methods

This study was an observational prospective study.

In May 2019, university students of Universidad San Francisco de Quito, in Quito, Ecuador, a university in an affluent region of Quito, were recruited. Participants were potentially eligible students who were on campus on two recruitment days during May 2019, and they were offered $US 10 in exchange for their participation. If they met eligibility criteria and agreed, informed consent was obtained. The study was previously approved by the USFQ (Universidad de San Francisco) Ethics committee on September 25th 2018 (2018-193IN).The inclusion and exclusion criteria are detailed in table 1. (***¡Error! No se encuentra el origen de la referencia***.)

**Table 1.** Inclusion and Exclusion Criteria for the participants in the study.

| Inclusion Criteria | Exclusion Criteria |
| --- | --- |
| • Aged 18-25 at the time of consent.<br>• Having suffered acne at any time point, including having active acne at the time of the study.<br>• Being a student of Universidad San Francisco de Quito USFQ.<br>• In case of using makeup during data collection, willing to remove it from the face to enable high quality photographs of the skin to be captured. | • Not fulfilling the inclusion criteria |

Once enrolled in the study, participants completed a survey that was developed based on the seventeen risk factors identified by Tan et al. and included questions addressing potential risk factors specific to the Ecuadorian population as determined by means of a Delphi expert consensus consulting registered Ecuadorian dermatologists. Each participant entered the photobooth and had three facial photographs taken following the standards stated in the pilot study publication^8^. The photographs of each participant were evaluated by three dermatologists who fulfilled a set of inclusion and exclusion criteria to participate in the study. Some of the most relevant inclusion criteria for the reviewing dermatologists were that they had practiced dermatology for more than five years, that they had seen more than 35 patients with acne in the last year, and that they had at least one publication related to acne in a scientific journal.

More details about the methods can be found in the publication of the pilot study that has been previously published^8^.

A necessary sample size was estimated to be around 250 participants, based on assumption of a recommended a minimum of 100 events and 100 non-events for logistic regression^9^, and an estimation from the literature that around 40% who go to a dermatologist have scars due to acne^4^.

In order to reduce the risk of information bias, data was collected on standardized forms as described in the study protocol and the participants simply responded to the survey described in Tan et al, 2020 on their own without any prompting or additional questions asked.

### Study variables

Study variables included: demographic variables (participant age, sex, ethnicity, region of residence), acne-related variables (acne severity (according to the participants themselves and the dermatologists), family history of acne scarring, age when acne first appeared, location of acne (face, neck, back, chest), duration of acne, squeezing and picking behaviors, acne scars (according to the participants themselves and the dermatologists), other types of scars, over the counter acne treatments, consultation of a dermatologist, prescribed acne treatments, if prescribed treatment was completed, treatment results), lifestyle behaviors (smoking status, fat diet), physical characteristics (weight, oily skin), and psychological or emotional stress.

A detailed explanation of self-assessments of acne scarring can be found in our previous publication.^8^

The outcome variable was the presence of acne scarring as determined by majority consensus of the 3 dermatologists.

### Statistical methods

For descriptive statistics, mean and standard deviation (std) were calculated for continuous variables and count and percentages for categorical variables. To determine risk factors and their respective weighting, logistic regressions were performed to estimate odds ratios (ORs) with 95% confidence intervals (95%CIs).. Univariate logistic models were performed to evaluate the association between each variable and the presence of acne scarring. A multivariate logistic regression model was then used with variables selected by clinical relevance and missing data rates. A p-value under 0.05 was considered statistically significant. All analyses were performed using SAS® version 9.4 (SAS Institute Inc., Cary, NC, USA).

## Results

404 participants signed informed consent and completed the survey to determine possible Ecuadorian risk factors and had 3 pictures taken. The mean patient age was 20·4 years with a standard deviation (std) of 1·8 (cf. Table 2). From the 404 patients, 63·6% were female and 36·4% were male. Concerning ethnicity, there were 87·4% mestizos and the rest were indigenous (7·0%), white (3.0%) and afro Ecuadorian (1·7%) and other (1%). Regarding the Ecuadorian region where the participants lived during their worst episodes of acne, the majority, 83·4% of patients lived in the Sierra (mountainous region). The rest of the participants lived in the Coastal region (10·9%), Amazonia (2 ·5%) and Galápagos Islands (0·25%).

**Table 2.** Demographic data and risk factors of the participants of the study.

|  |  | All patients (N=404) |
| --- | --- | --- |
| **Age at baseline** | Missing data | 4 |
|  | Mean (std) | 20.36 (1.82) |
| **Sex** | Missing data | 0 |
|  | Female | 257 (63.63%) |
|  | Male | 147 (36.39%) |
| **Ethnicity** | Missing data | 0 |
|  | Mestizo | 353 (87.38%) |
|  | Indigenous | 28 (6.93%) |
|  | White | 12 (2.97%) |
|  | Afro-Ecuadorian | 7 (1.73%) |
|  | Other | 4 (0.99%) |
| **Region** | Missing data | 0 |
|  | Sierra | 337 (83.42%) |
|  | Coast | 44 (10.89%) |
|  | Amazonia | 10 (2.48%) |
|  | Galápagos | 1 (0.25%) |
|  | Other | 12 (2.97%) |
| **Severity of acne (based on pictures)** | Missing data | 4 |
|  | A | 195 (48.75%) |
|  | B | 123 (30.75%) |
|  | C | 57 (14.25%) |
|  | D | 15 (3.75%) |
|  | E | 10 (2.50%) |
| **Age at the beginning of acne** | Missing data | 1 |
|  | Mean (std) | 14.78 (2.05) |
| **Duration of acne (years)** | Missing data | 28 |
|  | Mean (std) | 2.12 (2.00) |
| **Squeezing and picking behaviors** | Missing data | 1 |
|  | Never | 11 (2.73%) |
|  | Rarely | 53 (13.15%) |
|  | Sometimes | 114 (28.29%) |
|  | Frequently | 152 (37.72%) |
|  | All the time | 73 (18.11%) |
| **Acne scars (patient perspective)** | Missing data | 8 |
|  | No | 202 (51.01%) |
|  | Yes | 194 (48.99%) |
| **Scars from other causes (patients perspective)** | Missing data | 4 |
|  | No | 191 (47.75%) |
|  | Yes | 209 (52.25%) |
| **Treatment for acne (without prescription)** | Missing data | 1 |
|  | No | 174 (43.18%) |
|  | Yes | 229 (56.82%) |
| **Consultation of a dermatologist** | Missing data | 3 |
|  | No | 130 (32.42%) |
|  | Yes | 271 (67.58%) |
| **Use of prescribed treatment for acne** | Missing data | 18 |
|  | No | 40 (14.39%) |
|  | Yes | 238 (85.61%) |
| **Prescribed treatment for acne completed** | Missing data | 8 |
|  | No | 79 (32.38%) |
|  | Yes | 145 (59.43%) |
|  | Continue treatment | 20 (8.20%) |
| **Result of prescribed treatment** | Missing data | 60 |
|  | Disappeared | 89 (30.80%) |
|  | They improved, but didn't disappear completely | 175 (60.55%) |
|  | They didn't improve (no change) | 25 (8.65%) |
| **Smoking status** | Missing data | 0 |
|  | No | 342 (84.65%) |
|  | Yes | 62 (15.35%) |
| **Weight** | Missing data | 2 |
|  | Obese | 38 (9.45%) |
|  | Normal weight | 364 (90.55%) |
| **Oily skin** | Missing data | 0 |
|  | No | 108 (26.73%) |
|  | Yes | 296 (73.27%) |
| **Fatty diet** | Missing data | 0 |
|  | No | 132 (32.67%) |
|  | Yes | 272 (67.33%) |
| **Psychological or emotional stress** | Missing data | 0 |
|  | No | 110 (27.23%) |
|  | Yes | 294 (72.77%) |
| **Presence of acne: Face** | Missing data | 4 |
|  | No | 6 (1.50%) |
|  | Yes | 394 (98.5%) |
| **Presence of acne: Neck** | Missing data | 65 |
|  | No | 246 (72.57%) |
|  | Yes | 93 (27.43%) |
| **Presence of acne: Back** | Missing data | 21 |
|  | No | 107 (27.94%) |
|  | Yes | 276 (72.06%) |
| **Presence of acne: Chest** | Missing data | 47 |
|  | No | 190 (53.22%) |
|  | Yes | 167 (46.78%) |

58·3% of patients reported that they had a family history of acne. The mean age of the participants at the beginning of their acne was 14·8 years old with a standard deviation of 2·1; and the duration of the acne was 2·1 years with a standard deviation of 2·0. When it came to squeezing and picking behaviours, 18·1% did it all the time, 37·7% did it frequently and only 2·7% never picked or squeezed their lesions.

56·82% of the participants reported to have used over-the-counter treatments such as soaps, creams or pills, and the majority (67·6%) reported that they consulted a dermatologist during their worst acne breakout. It is of note that 85·6% of the participants used a treatment prescribed by a specialist; but only 59·4% of the patients fully completed the prescribed treatment. Descriptive analyses of all potential risk factors considered can be found in table 2.

Results from univariate analyses indicated that male sex (OR=2.76 95%CI [1.72; 4.43]), severe or very severe acne scarring (OR=4.28 95%CI [1.24; 14.79]), acne duration over 1 year (OR=1.71 95%CI [1.12; 2.60]), oily skin (OR=2.02 95%CI [1.27; 3.22]) and the presence of acne on the neck (OR=2.26 95%CI [1.30; 3.92]), were all significantly associated with the presence of acne scarring (cf. Table 3). According to the multivariate regression, male sex (2.56 95%CI [1.58;4.17]), oily skin (1.96 95%CI [1.20;3.20]) and severe or very severe acne (3.7595%CI [1.05;13.37]) remained significant risk factors for acne scarring. Duration of acne was kept in the model considering its universally and consistently known clinical importance and impact on acne scarring (cf. Table 4).

**Table 3.** Univariate analysis of the risk factors for acne scarring.

|  | N | OR [95% CI] | P-value |
| --- | --- | --- | --- |
| Age at baseline | 366 | 1.01 [0.90;1.14] | 0.8679 |
| Sex (male vs female) | 370 | 2.76 [1.72;4.43] | <0.0001 |
| Severity of acne (severe- very severe vs almost clear - moderate) | 367 | 4.28 [1.24;14.79] | 0.0217 |
| Severity of acne - bis (Moderate - severe - very severe vs Almost clear) | 367 | 2.51 [1.63;3.85] | <0.0001 |
| Family history of acne scarring | 369 | 1.02 [0.67;1.56] | 0.9230 |
| Age at the beginning of acne | 369 | 1.01 [0.91;1.12] | 0.8068 |
| Duration of acne (>1 year vs <=1 year) | 370 | 1.71 [1.12;2.60] | 0.0126 |
| Duration of acne (>2 years vs <=2 years) | 370 | 1.89 [1.16;3.06] | 0.0102 |
| Duration of acne (>3 years vs <=3 years) | 370 | 1.49 [0.84;2.66] | 0.1744 |
| Squeezing and picking behaviors (Frequently - all the time vs Never - sometimes) | 369 | 0.86 [0.56;1.31] | 0.4758 |
| Scars from other causes (patient perspective) | 366 | 0.79 [0.52;1.20] | 0.2616 |
| Treatment for acne (without prescription) | 369 | 0.83 [0.54;1.26] | 0.3751 |
| Smoking status | 370 | 0.98 [0.55;1.75] | 0.9531 |
| Weight (Obese vs Normal weight) | 368 | 0.68 [0.34;1.36] | 0.2756 |
| Oily skin | 370 | 2.02 [1.27;3.22] | 0.0029 |
| Fatty diet | 370 | 1.09 [0.70;1.69] | 0.7174 |
| Psychological or emotional stress | 370 | 1.05 [0.66;1.66] | 0.8495 |
| Presence of acne: Neck | 308 | 2.26 [1.30;3.92] | 0.0039 |
| Presence of acne: Back | 351 | 1.20 [0.75;1.92] | 0.4523 |
| Presence of acne: Chest | 326 | 1.25 [0.80;1.95] | 0.3297 |

**Table 4.** Multivariate analysis of the risk factors for acne scarring.

|  | OR [95%CI] | P-value |
| --- | --- | --- |
| Sex (Male vs Female) | 2.56 [1.58;4.17] | 0.0001 |
| Duration of acne (>1 year vs <=1 year) | 1.38 [0.88;2.15] | 0.1581 |
| Oily skin | 1.96 [1.20;3.20] | 0.0067 |
| Severity of acne (Severe - very severe vs Almost clear - moderate) | 3.75 [1.05;13.37] | 0.0419 |

## Discussion and conclusion

Our study found that in the Ecuadorian population, the most statistically significant risk factors associated with acne scarring included male sex, severe or very severe acne, duration of acne (as this is universally known to have a clinically important impact), the presence of oily skin and the presence of acne on the neck. These are slightly different from the ones determined for the scar risk assessment tool described in Tan et al. which were severity of acne, family history of acne scarring, duration of acne and squeezing and picking behaviors. Even though squeezing and picking behaviors and family history of acne scarring risk factors were classified as most relevant in Tan et al.’s study, our study found that male sex, the presence of oily skin and the presence of acne in the neck were the most important factors associated with acne scarring. We understand that these differences may be due to various factors and that the risk factors identified in Tan et al cannot be disregarded. Considering the risk factors identified in our study, we believe the presence of acne in the neck being a risk factor may be related to the fact that this country is located in the equatorial line where sun exposure is direct and stronger than in other countries. In the case of oily skin, it must be noted that of the largest population in this study was of mestizo ethnicity; thus there may be a relationship between race and a certain type of skin that should be further analyzed and has not been previously explored. Finally, male sex being a risk factor can be a result of many underlying issues. First greater association between male sex with acne scarring that we found could be related to direct sun exposure and additionally the fact that men are less prone to apply sunscreen than women, as supported by the participating dermatologists.

### Limitations

One of the shortcomings of our study is that the participants were recruited at Universidad San Francisco de Quito, therefore the majority belonged to the mountainous region (Sierra) of Ecuador, which could lead to a selection bias. Additionally, this university is in a highly affluent section of Quito, Ecuador, a large metropolitan region. Therefore, we understand that this presents an important selection bias. The generalizability of our results may be limited and further studies need to be conducted to confirm our findings.

There could be a small degree of recall bias as participants had to use their memory to determine when they were first diagnosed with acne and the duration of their acne. However, we do not feel this bias as important considering most of the participants were not far from their adolescence and had no particular preconceptions about risk factors for acne scarring. Nonetheless, when the participant could not remember their acne history, missing data happened, such as for the duration of acne.

We acknowledge that there were some discrepancies across dermatologists regarding what was considered acne scarring and the degree of acne scarring. We tried to limit the impact of this heterogeneity by asking several dermatologists’ opinion and decide the result using the majority. A pilot study conducted prior to this study suggested that this method using three local dermatologists corresponds to an adequate proxy for international experts’ evaluation of individual acne scarring.

### Conclusions

By identifying acne scarring risk factors and applying the tool in everyday dermatology consults, we can reduce the physical and psychological burden that acne scarring causes in the adolescent and adult population. Further research should be performed in order to reassess potential risk factors and complete the adaptation of the tool for the Ecuadorian population, with a larger and more representative study population. Other researchers in different countries or geographical regions should also consider adapting such tools to their populations due to the fact that race, demographics and climate can be determinants of acne scarring risk factors, among others.

## Data Availability

Harvard Dataverse: Questionnaire for the validation and adaptation of a tool to estimate the risk of acne-induced scars in different
population. https://doi.org/10.7910/DVN/WGDWB015.
Harvard Dataverse: Protocol for the validation and adaptation of a tool to estimate the risk of acne-induced scars in different populations. https://doi.org/10.7910/DVN/NED0GS16
Extended data are available under the terms of the Creative Commons Zero No rights reserved data waiver (CC0 1.0 Public domain dedication)

https://doi.org/10.7910/DVN/WGDWB015

## References

1. Oge’, L. K., Broussard, A., & Marshall, M. D. (2019). Acne Vulgaris: Diagnosis and Treatment. American Family Physician, 100(8), 475–484.

2. Sing, Anna H., & Chew Fook T. (2020). Systematic review of the epidemiology of acne vulgaris. Scientific Reports, 10(5754), 29. https://doi.org/10.1038/s41598-020-62715-3

3. Layton, A. M., Thiboutot, D., & Tan, J. (2021). Reviewing the global burden of acne: How could we improve care to reduce the burden?*. British Journal of Dermatology, 184(2), 219–225. https://doi.org/10.1111/bjd.19477

4. Tan, Jerry & Leyden, James. (2017, febrero 1). Prevalence and Risk Factors of Acne Scarring Among Patients Consulting Dermatologists in the USA. JDDonline - Journal of Drugs in Dermatology. https://jddonline.com/articles

5. Habeshian, K. A., & Cohen, B. A. (2020). Current Issues in the Treatment of Acne Vulgaris. Pediatrics, 145(Supplement 2), S225–S230. https://doi.org/10.1542/peds.2019-2056L

6. Bhargava, S., Cunha, P. R., Lee, J., & Kroumpouzos, G. (2018). Acne Scarring Management: Systematic Review and Evaluation of the Evidence. American Journal of Clinical Dermatology, 19(4), 459–477. https://doi.org/10.1007/s40257-018-0358-5

7. Tan, J., Thiboutot, D., Gollnick, H., Kang, S., Layton, A., Leyden, J. J., Torres, V., Guillemot, J., & Dréno, B. (2017). Development of an atrophic acne scar risk assessment tool. Journal of the European Academy of Dermatology and Venereology: JEADV, 31(9), 1547–1554. https://doi.org/10.1111/jdv.14325

8. Estrella Porter, J., Camacho, M., Viteri, M. I., Aguilar, K., Belhadi, D., Bettoli, V., Buestán, A. del R., Dréno, B., Endara, P., Layton, A., Machado, N., Mateus, R., Tan, J., Terán, E., Yépez, P., & Guillemot, J. (2020). Pilot study for the evaluation and adaptation of a Four Item-Acne-Scar Risk Assessment Tool (4-ASRAT): A resource to estimate the risk of acne-induced scars. F1000Research, 9, 651. https://doi.org/10.12688/f1000research.23737.1

9. “Substantial effective sample sizes were required for external validation studies of predictive logistic regression models.” Journal of clinical epidemiology 58.5 (2005): 475–483.

